# Long-term Efficacy of Compound Trabeculectomy in the Treatment of Uveitic Glaucoma: A Retrospective Cohort Study

**DOI:** 10.64898/2026.02.06.26345693

**Authors:** Xinyue Ji, Xinmiao Shan, Long fang Zhou, Lili Jing, Xiaoxi Liu, Jiajun Wei, Xiaojing Pan, Die Hu

**Author notes:** **Correspondence Author:** Xiaojing Pan, MD, PhD, Eye Institute of Shandong First Medical University, Qingdao Eye Hospital of Shandong First Medical University, 5 Yanerdao Road, Shinan District, Qingdao 266071, Shandong Province, China, Die Hu, MM, Eye Institute of Shandong First Medical University, Qingdao Eye Hospital of Shandong First Medical University, 5 Yanerdao Road, Shinan District, Qingdao 266071, Shandong Province, China. Dr Ji and Shan contributed equally as co-first authors. Dr Pan and Hu contributed equally as co-last authors.

## Abstract

**Purpose:** To evaluate the three-year efficacy and safety of compound trabeculectomy for uveitic glaucoma (UG).

**Methods:** This retrospective study enrolled 51 patients (53 eyes) requiring compound trabeculectomy, divided into UG (25 eyes) and non-UG groups (28 eyes). Outcomes including intraocular pressure (IOP), medication use, surgical success rates, and complications were analyzed over 3 years.

**Results:** Baseline characteristics including age, sex, preoperative IOP and medication use were comparable (all P>0.05). At 36 months, postoperative IOP was showed no significant differences, which was 15.4±8.4 mmHg and 14.6±3.3 mmHg (P=0.73) with 54% and 55% reduction (P=0.88) in UG and non-UG groups respectively. The qualified success rate was 76.0% and 85.7% at 36 months in UG and non-UG group, and Kaplan–Meier analysis showed no significant difference. Medication reduction of UG group was significant lower than non-UG group (P=0.0058). Comparable complication rates were observed between groups (all P>0.05), yet bleb scarring and cataract progression showed elevated incidence in both cohort.

**Conclusion:** Compound trabeculectomy effectively reduced IOP and medications use in UG and non-UG. There was no significant difference in both qualified and completed success rate between UG and non-UG. Complications of filtering bleb fibrosis and cataract progression should be pay close attention for both groups.

## Introduction

Uveitis, an inflammatory disorder of the uveal tract, is anatomically classified into anterior, intermediate, posterior, and panuveitis according to the International Uveitis Study Group classification system (SUN) (Bloch-Michel E et al, 1987; Massa H et al, 2019; Sudharshan S et al, 2010). Anterior uveitis predominates in clinical practice, accounting for 50% of uveitis cases in China (Yanoff et al, 2009). Notably, the incidence of viral anterior uveitis (VAU) has exhibited a progressive annual increase (Yang et al, 2017), and VAU characterized by persistent ocular hypertension is particularly prevalent in coastal regions (Pan, 2020). Uveitis represents a leading cause of preventable blindness, frequently resulting in severe complications such as glaucoma, cataracts, macular edema, and retinopathy (Pillai MR et al, 2024; Škrlová E et al, 2023). About 20% of patients with chronic anterior uveitis will develop secondary glaucoma, a particularly vision-threatening complication (Škrlová et al., 2023). Untreated cases progress to irreversible optic neuropathy, with blindness prevalence exceeding 30% (Nema et al., 2018; Rojas-Carabali et al., 2024). Therefore, uveitic glaucoma (UG) management requires prioritized inflammatory control followed by aggressive intraocular pressure regulation.

Current therapeutic strategies for uveitic glaucoma include medical, laser, and surgical interventions. In the case of persistent elevation of IOP despite infIammation reduction, and in the case of the optic nerve damage, it is appropriate to proceed to anti-glaucoma surgery such as trabeculectomy (Škrlová E et al, 2023; Koktekir et al., 2013; Ma et al., 2022). The gold standard of anti-glaucoma procedure is trabeculectomy, demonstrated broad clinical applicability particularly in patients unresponsive to pharmacological or laser interventions (Modi et al., 2011; Škrlová et al., 2023; Urbonavičiūtė et al., 2022). During development of several decades, in order to decrease a series of complications such as shallow anterior chamber and filtering bleb fibrosis, modern surgical advancements have introduced critical innovations, including the removable suture technique, anti-metabolic drugs (e.g., mitomycin C), anterior chamber puncture technique, intraoperative pre-evaluation of postoperative excessive filtration, and postoperative filtration bubble massage (Glaucoma Group of Chinese Ophthalmological Society, 2017; Wang et al, 2023; Gao Y et al, 2022). This modified trabeculectomy techniques is called “compound trabeculectomy”. These enhancements of compound trabeculectomy collectively improve surgical efficacy while mitigating complications, resulting in significantly higher success rates compared to conventional trabeculectomy (Glaucoma Group of Chinese Ophthalmological Society, 2017; Wang et al, 2023; Gao Y et al, 2022; Yuan et al, 2016).

Some researchers studied the outcomes of trabeculectomy compared UG with POAG, and there was no significent difference of the success rate. Kanaya et al. (2021) reported that the success rates (IOP<18 mmHg, IOP reduction >20%) in UG and POAG were 82.2% and 75.6% at 36 months respectively. Kaburaki et al. (2009) reported the comparable complete success rate (≤15 mmHg) for postoperative IOP control at 5 years were 57.1% and 53.7% in UG and in POAG respectively. But Iwao et al. (2014) demonstrated that trabeculectomy was less effective in maintaining IOP reduction in UG eyes than in POAG eyes, with 71.3% and 89.7% of the 3-year probabilities of success, respectively. However, the trabeculectomy in those three studies were traditional trabeculectomy or with one or two surgical advancements. After the introduction and development of compound trabeculectomy, a few researches studied the its effciency as PACG or POAG treatment, mainly. Gao et al. (2022) investigate the efficacy of peripheral iridectomy and compound trabeculectomy for primary angle-closure glaucoma. Yuan et al. (2016) reported that compound trabeculectomy can effectively control the intraocular pressure of APACG patients and decrease the incident rate of postoperative complications. Wang et al. (2023) demonstrated that compound trabeculectomy appears to be an effective treatment option for secondary glaucoma when IOP cannot be controlled through topical medications. Neverthless, few studies have assessed the outcomes to control IOP and decrease complications of compound trabeculectomy for UG compared with non-UG, in a strict sense. As the inheritance and innovation of the gold standard, we should evaluate the efficacy and safety of compound trabeculetomy for patients with uveitic secondary glaucoma once again.

This three-year prospective, single-center and non-random comparative study evaluates long-term surgical outcomes between uveitic and non-uveitic glaucoma cohorts who were undergo compound trabeculectomy, aiming to evaluate the efficacy and safety of compound trabeculetomy and establish evidence-based safety and efficacy parameters for optimizing surgical protocols in uveitic glaucoma.

## Method

### Study Design

The study was retrospective. Patients were consecutively enrolled from August 2020 to April 2022 at the Eye Institute of Shandong First Medical University, Qingdao Eye Hospital of Shandong First Medical University, China. We collected 51 patients of UG and other types of glaucoma who needed to conducted compound trabeculacetomy, and followed up for 3 years to collect their IOP, medications, surgery qualified success rate and complete success rate as well as complications, to assess the safety and effiency of the compound trabeculectomy. The study adhered to the tenets of the Declaration of Helsinki and was approved by the Ethics Committee of Qingdao Eye Hospital, affiliated to Shandong First Medical University ((Approval No.: Qingyan Lunshen (Fast) [2026] 14)). Informed consent was obtained from each participant prior to the procedures. Any patient identifiers haven’t be used to identify individuals.

### Patients Selection

Patients with UG were enrolled according to the following criteria: a) recurrent mild anterior uveitis; b) elevated IOP (>21 mmHg) that were difficult to control even with the maximum dosage of medication; c) various shapes of keratic precipitates (fine, focal, mutton-fat or stellate); d) no iris posterior synechiae or peripheral anterior synechiae; e) with mild or without iris stromal atrophy; f) with mild or without posterior inflammation; g)a minimum follow-up of 36 months. The non-UG patients were enrolled if they met the following criteria: a) no uveitis; b) elevated IOP (>21 mmHg) that were difficult to control even with the maximum dosage of medication; c).a minimum follow-up of 36 months.

All uveitis patients were treated with corticosteroids eye drops in pre-operation. If necessary, we used antiviral eye drops to control inflammation until inflammation cells ≤0.5+. After compound trabeculectomy, all patients would use corticosteroids and nonsteroidal eye drops for one month to control inflammation.

### Surgery Treatment

All patients underwent compound trabeculectomy, which was performed by a same experienced surgeon. All surgery procedures were under peribulbar anesthesia with lidocaine. The surgical procedure is as follows: Surgical procedures were initiated with standardized preparation of a conjunctival-scleral flap (3 mm × 4 mm, half-thickness) anchored at the corneal limbus. Antifibrotic agents (mitomycin C: 0.3 mg/mL or 5-fluorouracil: 25–50 mg/mL) were applied intraoperatively for 1-2 min to inhibit fibrosis, followed by saline irrigation. Anterior chamber paracentesis preceded excision of a 2 mm × 2 mm corneoscleral block with peripheral iridectomy to establish aqueous drainage. The scleral flap was secured using 10-0 nylon interrupted sutures and adjustable releasable sutures (corneal fixation) to modulate filtration. Bleb functionality was verified via balanced saline infusion, with continuous conjunctival closure and anterior chamber stabilization ensuring structural integrity.

### Data Analysis

Preoperative data included surgical age, sex, diagnosis etiology, and baseline medication count. Three-year postoperative outcomes compared between the groups comprised IOP, medication use, success rates (qualified/complete), and complications. Surgical success was categorized as complete (postoperative IOP ≤21 mmHg with ≥20% reduction without medications) or qualified (postoperative IOP ≤21 mmHg with ≥20% reduction requiring medications).

Statistical analyses were conducted using R software version 4.4.2. Continuous variables conforming to normal distribution (assessed via Shapiro-Wilk test) and homogeneity of variance (Bartlett’s test) were presented as mean ± standard deviation (SD) and analyzed with Student’s t-test for between-group comparisons. When parametric assumptions were violated, the Mann-Whitney U test (Wilcoxon rank-sum test) was substituted. Multi-timepoint comparisons were analyzed using repeated-measures Analysis of Variance (ANOVA) with Mauchly’s sphericity test, with Greenhouse-Geisser correction and Bonferroni-adjusted post hoc tests applied when sphericity assumptions were violated. Categorical variables were reported as counts (percentages) and compared using Pearson’s chi-square test or Fisher’s exact test. Statistical significance was defined as a two-tailed p-value < 0.05.

## Result

### Baseline Characteristics

Table 1 summarizes the baseline characteristics of the study cohort comprising 23 uveitis patients (25 eyes) and 26 non-uveitis patients (28 eyes). The uveitis group demonstrated anterior uveitis (15 eyes [60.0%]) and Fuchs syndrome (7 eyes [28.0%]) as predominant diagnoses, while the non-uveitis group showed primary open-angle glaucoma (12 eyes [42.9%]) and primary angle-closure glaucoma (5 eyes [17.9%]). No significant differences were observed in surgical age (uveitis: 53.0±9.2 years vs non-uveitis: 57.0±19.2 years; P=0.132), gender distribution (male: 65.2% vs 61.5%; P=1.000), or preoperative glaucoma medications (uveitis: 2.96±1.34 vs non-uveitis: 2.43±1.40; P=0.165).

**Table 1.** Baseline Characteristics and diagnosis.

|  | Uveitis Group<br>n=25 | Non Uveitis Group<br>n=28 | P Value |
| --- | --- | --- | --- |
| **Age\*** | 53.04 ± 9.19 | 57.00 ± 19.18 | 0.132 |
| **Sex\*** |  |  | 1.000 |
| Male | 15 (65.2%) | 16 (61.5%) |  |
| Female | 8 (34.8%) | 10 (38.5%) |  |
| **IOP (mmHg)** | 33.61± 9.29 | 32.35± 12.23 | 0.676 |
| **Medication** | 2.96 ±1.34 | 2.43 ± 1.40 | 0.165 |
| **Uveitis etiology** |  |  | — |
| Anterior uveitis | 15 (60.0%) | — |  |
| Fuchs Syndrome | 7 (28.0%) | — |  |
| Herpes virus uveitis | 1 (4.0%) | — |  |
| Posner-Schlossman<br>Syndrome | 1 (4.0%) | — |  |
| Behcet disease | 1 (4.0%) | — |  |
| A-R Syndrome | — | 2 (7.1%) |  |
| Neovascular glaucoma | — | 2 (7.1%) |  |
| Glucocorticoid induced<br>glaucoma | — | 2 (7.1%) |  |
| Primary open-angle<br>glaucoma | — | 12 (42.9%) |  |
| Primary angle closed<br>glaucoma | — | 5 (17.9%) |  |
| UGH Syndrome | — | 2 (7.1%) |  |
| Traumatic Secondary<br>glaucoma | — | 3 (10.8%) |  |

Data are presented as mean (± SD) and frequencies (%). Numbers are per eyes.
\*Numbers are per patients.

### IOP

Baseline and follow-up IOP for both groups are detailed in Table 2 and visualized in Figures 1-2a. Initial IOP measurements showed comparable preoperative values between uveitis (33.6±9.3 mmHg) and non-uveitis groups (32.4±12.2 mmHg). Mauchly’s test revealed significant sphericity violations (Time: W=0.038, p=1.38×10□^23^; Interaction: W=0.038, p=1.38×10□^23^), necessitating Greenhouse-Geisser correction. Repeated measures ANOVA demonstrated significant temporal effects (F[2.76,140.65]=78.717, p=1.45×10□^2^□) without intergroup differences (Group: F[1,51]=0.014, p=0.905; Interaction: F[2.76,140.65]=0.476, p=0.684). Bonferroni-adjusted pairwise comparisons confirmed preoperative IOP exceeded all postoperative measurements (17.7-20.5 mmHg; all p<0.0001), while inter-postoperative intervals showed no significant variations (all p>0.05 vs baseline). Three-year outcomes revealed comparable IOP levels (uveitis:15.4±8.4 mmHg vs non-uveitis:14.6±3.3 mmHg; P=0.656) and equivalent percentage reductions from baseline (54% vs 55%; P=0.880), with all longitudinal comparisons remaining non-significant at adjusted α=0.0024.

**Table 2.**
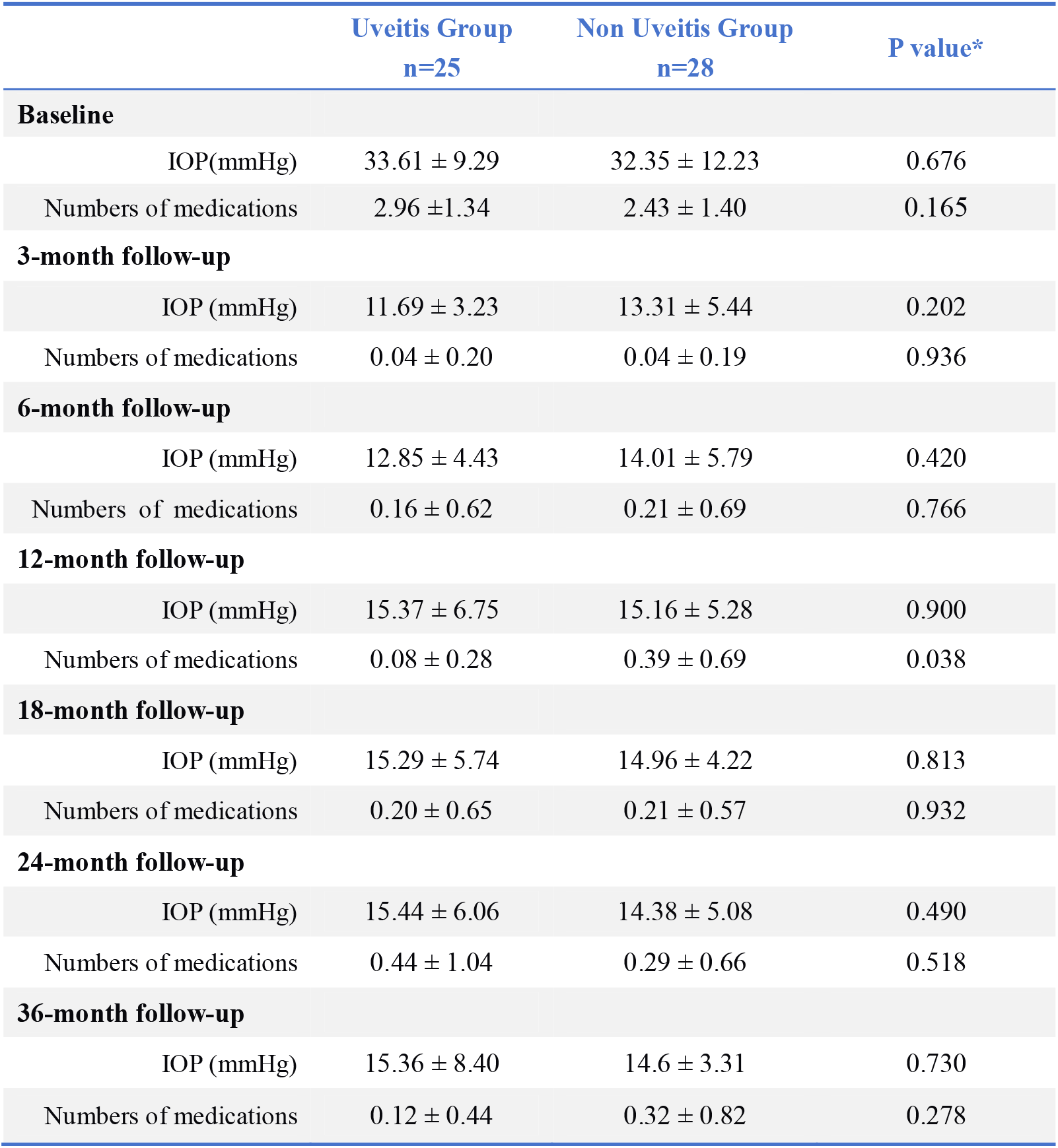
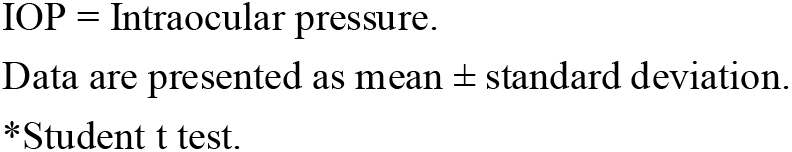
Intraocular Pressure and Medical Therapy at Baseline and Follow-up.

|  | **Uveitis Group**<br>**n=25** | **Non Uveitis Group**<br>**n=28** | **P value\*** |
| --- | --- | --- | --- |
| **Baseline** |  |  |  |
| IOP(mmHg) | $33.61 \pm 9.29$ | $32.35 \pm 12.23$ | 0.676 |
| Numbers of medications | $2.96 \pm 1.34$ | $2.43 \pm 1.40$ | 0.165 |
| **3-month follow-up** |  |  |  |
| IOP (mmHg) | $11.69 \pm 3.23$ | $13.31 \pm 5.44$ | 0.202 |
| Numbers of medications | $0.04 \pm 0.20$ | $0.04 \pm 0.19$ | 0.936 |
| **6-month follow-up** |  |  |  |
| IOP (mmHg) | $12.85 \pm 4.43$ | $14.01 \pm 5.79$ | 0.420 |
| Numbers of medications | $0.16 \pm 0.62$ | $0.21 \pm 0.69$ | 0.766 |
| **12-month follow-up** |  |  |  |
| IOP (mmHg) | $15.37 \pm 6.75$ | $15.16 \pm 5.28$ | 0.900 |
| Numbers of medications | $0.08 \pm 0.28$ | $0.39 \pm 0.69$ | 0.038 |
| **18-month follow-up** |  |  |  |
| IOP (mmHg) | $15.29 \pm 5.74$ | $14.96 \pm 4.22$ | 0.813 |
| Numbers of medications | $0.20 \pm 0.65$ | $0.21 \pm 0.57$ | 0.932 |
| **24-month follow-up** |  |  |  |
| IOP (mmHg) | $15.44 \pm 6.06$ | $14.38 \pm 5.08$ | 0.490 |
| Numbers of medications | $0.44 \pm 1.04$ | $0.29 \pm 0.66$ | 0.518 |
| **36-month follow-up** |  |  |  |
| IOP (mmHg) | $15.36 \pm 8.40$ | $14.6 \pm 3.31$ | 0.730 |
| Numbers of medications | $0.12 \pm 0.44$ | $0.32 \pm 0.82$ | 0.278 |

IOP = Intraocular pressure.
Data are presented as mean ± standard deviation.
\*Student t test.

**Figure 1.**
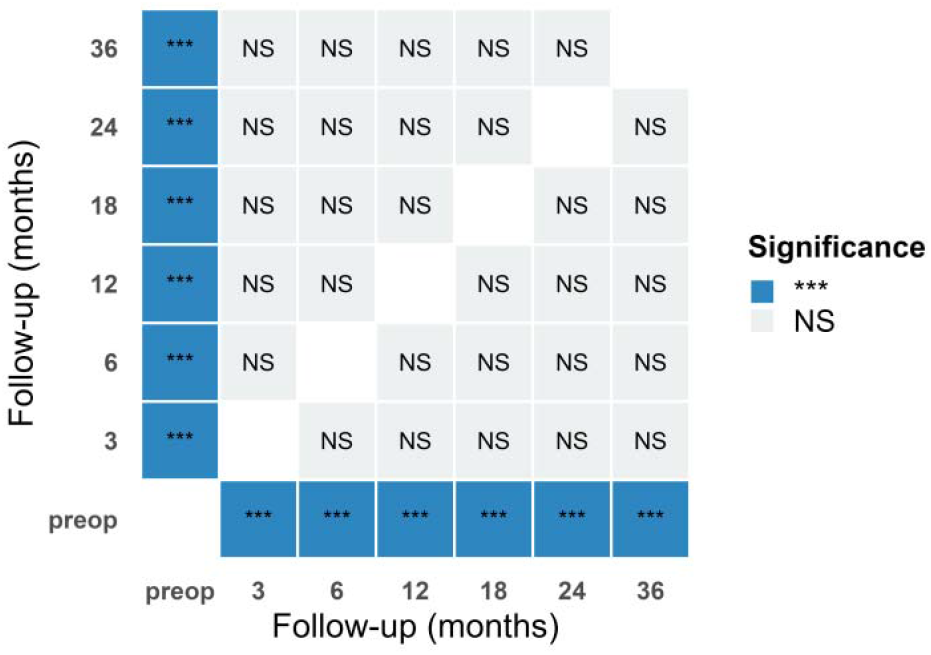
Significance of Different Time Point. *** = Significance; NS = No Significance.

**Figure 2.**
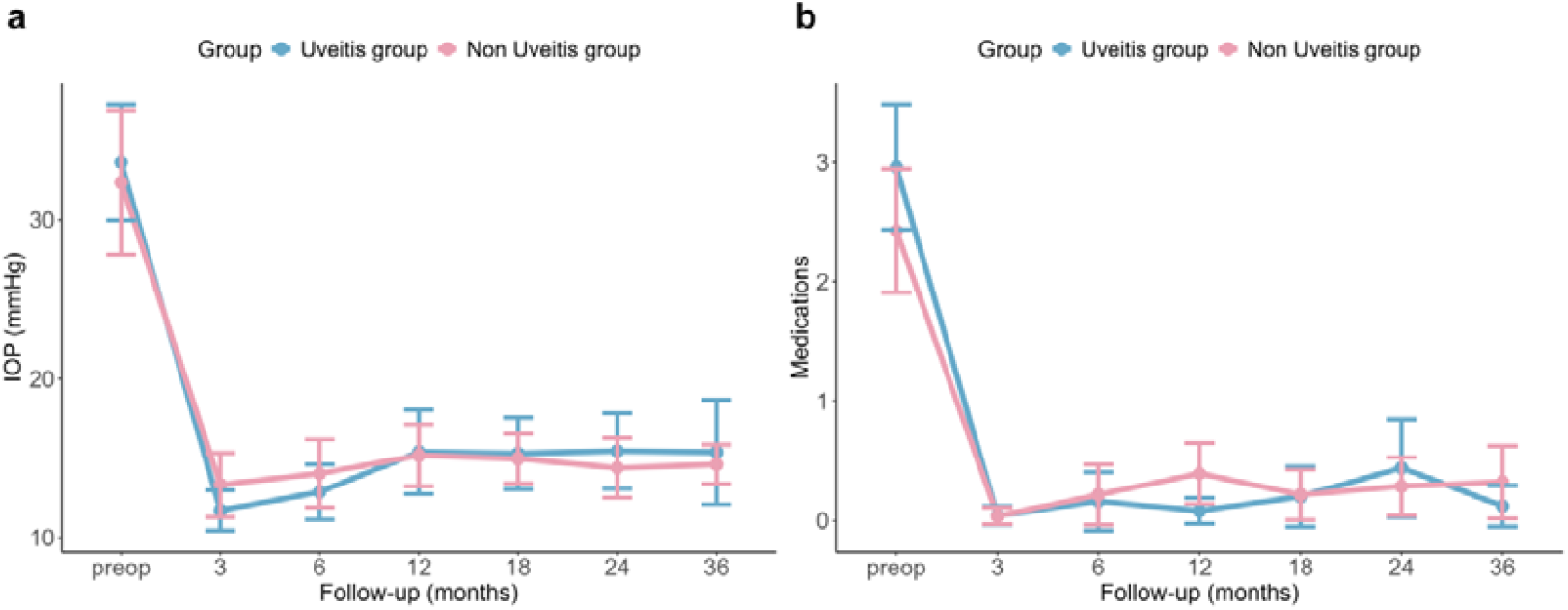
(a) IOP changes over time (mean±95%CI). (b) Medications changes over time (mean± 95%CI).

### Medical therapy

Medication use patterns are detailed in Table 2. Preoperative glaucoma medication counts showed no significant intergroup difference (uveitis: 2.96±1.34 vs non-uveitis: 2.43±1.40; P=0.165). Significant divergence emerged at 12-month evaluation (uveitis: 0.08±0.28 vs 0.39±0.69; P=0.038). At last follow-up time of 3 years, the numbers of medications showed no significance (0.12±0.44 vs 0.32±0.82; P=0.278). Medication reduction from baseline to 3 years was greater in uveitis patients (2.84±1.31 vs 2.11±1.42; P=0.0058). No other follow-up intervals demonstrated significant between-group medication variations.

### Surgery outcomes

Kaplan-Meier survival analysis with log-rank testing demonstrated comparable therapeutic outcomes between groups in Figure 3. Qualified success rates for the uveitis versus non-uveitis groups were 88.0% vs 89.3% at 12 months, 84.0% vs 85.7% at 24 months, and 76.0% vs 85.7% at 36 months (P=0.40). Complete success rates exhibited progressive declines: 96.0% at 3 months, 92.0% at 6 months, 80.0% at 12 months, 72.0% at 18 and 24 months, 68.0% at 36 months in the uveitis group, versus 96.4%, 89.3%, 64.3%, 60.7%, 53.6%, and 50.0% at corresponding intervals in the non-uveitis group (P=0.20).

**Figure 3.**
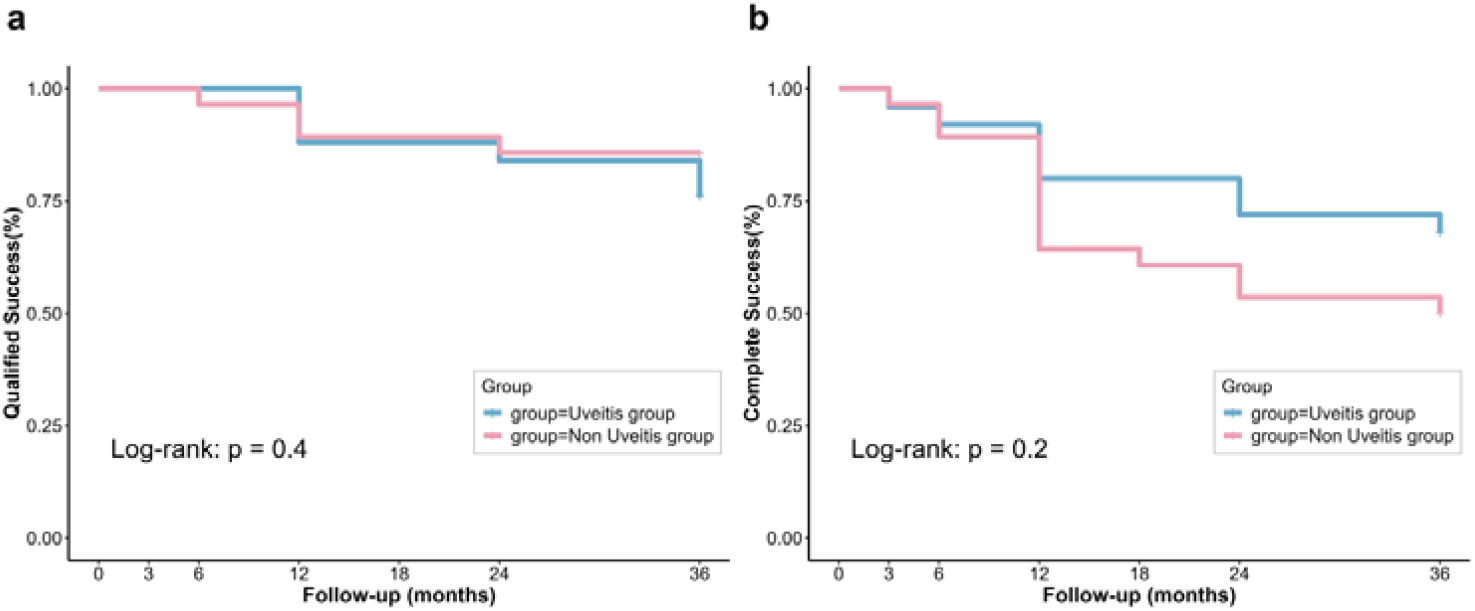
Kaplane-Meier plots showing the success rate in the uveitis group and non-uveitis group. (a). Qualified success (%) was defined as postoperative IOP between 6and 21 mmHg, and decreased at least 20% compared to the baseline IOP with or without anti-glaucoma medications. (b). Complete success (%) was the same as qualified success without anti-glaucoma medications.

### Complication

Postoperative complications are summarized in Table 3. Filtering bleb fibrosis constituted the predominant complication (uveitis:48.0% vs non-uveitis:67.9%; P=0.236, Pearson χ^2^ test), followed by cataract progression (36.0% vs 17.9%; P=0.237). Both cohorts exhibited comparable rates of hypotony with shallow anterior chamber (1 eye each; P=1.000, Fisher’s exact test) and bleb leakage (uveitis:2 vs non-uveitis:1; P=0.973). Choroidal detachment occurred in 1 uveitis patient versus 2 non-uveitis patients (P=1.000). Unique to the non-uveitis cohort were hyphema (1 eye) and malignant glaucoma (1 eye). Uveitis patients demonstrated anterior chamber inflammation recurrence in 46.2% cases, with 30.8% experiencing ≥2 episodes.

**Table 3.** Postoperative complications in the uveitis and non-uveitis groups.

|  | Uveitis Group<br>n=25 | Non Uveitis Group<br>n=28 | P Value |
| --- | --- | --- | --- |
| **Complicated cataract** | 9 (36.0%) | 5 (17.9%) | 0.237* |
| **Hypotony with shallow anterior chamber** | 1 (4.0%) | 1 (3.6%) | 1.000** |
| **Leaking bleb** | 2 (8.0%) | 1 (3.6%) | 0.973** |
| **Hyphema** | 0 | 1 (3.6%) | 1.000** |
| **Filtering bleb fibrosis** | 12 (48.0%) | 19 (67.9%) | 0.236* |
| **Malignant glaucoma** | 0 | 1 (3.6%) | 1.000** |
| **Choroidal detachment** | 1 (4.0%) | 2 (7.1%) | 1.000** |

Data presented as number of patients (percentage).
\*The Pearson chi-square test.
\*\*Fisher's test.

## Discussion

Trabeculectomy, first introduced by Cairns in 1968, remains the “gold standard” surgical intervention for glaucoma by creating an alternative aqueous humor drainage pathway through a conjunctival filtering bleb (Gao X et al., 2022; Zhang X et al., 2023). After several years development, trabeculectomy can be augmented with adjunctive techniques, including the removable suture technique, antimetabolite application (e.g., mitomycin C), anterior chamber paracentesis, intraoperative assessment of potential postoperative overfiltration, and postoperative bleb massage, which is called compound trabeculectomy (Glaucoma Group of Chinese Ophthalmological Society, 2017; Gao et al, 2022; Yuan et al, 2016; Wang et al, 2023). This modern procedure enhances success rates, significantly reduces fibrosis and minimizes early overfiltration complications (Weinreb et al.,2015; Weinreb et al.,2016; Zhou M et al.,2014)..

Our study observed maximal IOP reduction at postoperative 3 months (uveitis group:21.92 mmHg decrease vs non-uveitis:19.04 mmHg). IOP gradually increased between 3 to 12 months before stabilizing near 15 mmHg, maintaining this level through year 3. The possible reasons of this trendency may be related to Tenon’s capsule fibrosis progression: early inflammation (0-7 days) involves type III collagen deposition, transitioning to type I collagen production through fibroblast-myofibroblast conversion at 3 months, ultimately achieving scar maturation by 12 months (Feldman et al., 2013; Kwon et al, 2017). Addtionally, filtering bleb morphology also significantly influences intraocular pressure (IOP) regulation. Kojima et al. (2015) demonstrated progressive bleb wall thickening through postoperative 3 months, and structural maturation at 6 months with subsequent filtration opening narrowing between 6 to 12 months. Notably, the IOP change tendency in our two groups aligned with those fibrotic dynamics. Furthermore, the three-year follow-up in our study provided adequate time for the bleb scarring-related IOP changes to manifest distinctly and reach the stabilization eventually.

Comparative analysis demonstrated equivalent trabeculectomy success rates between uveitis and non-uveitis groups, which is 76.0% and 85.7% at 36 months in UG and non-UG groups. though previous study has reported the 71.3% and 89.7% success rate (<21 mmHg) of the 3-year probabilities of success in UG and POAG patients and indicated the success rate of trabeculectomy in the uveitis group is significantly lower than in other types of secondary glaucoma (Iwao et al, 2014). Our success rate of UG was slightly higher than Iwao et al. (2014) outcomes, and this phenomenon may attribute to the predominance of anterior uveitis in the uveitis cohort (n=15 eyes, 60% anterior uveitis). Meanwhile, some patients with refractory glaucoma were included in the non-UG group, leading to lower success rate than previous study (Iwao et al, 2014). For pathophysiology of most anterior uveitis patients in the UG group, inflammation primarily localized to the trabecular meshwork and corneal endothelium manifested as stellate keratic precipitates (KP), anterior chamber flare, and elevated intraocular pressure and this anterior segment-predominant inflammation exhibited mild overall severity (Babu K et al,2020; Yanoff et al. 2009). Based on the clinical manifestations of anterior uveitis patients, we speculate that the diagnosis of these patients is highly likely to be viral anterior uveitis. Therefore, the possible interpretations of the elevated surgical success rates observed in the uveitis cohort may be related to three principal factors. Firstly, inflammatory cells (e.g., neutrophils, lymphocytes) or viral factors in the anterior chamber are efficiently drained through the newly established filtration pathway (Yanoff et al.,2009; Sen H N et al, 2024). This mechanism achieves significant reductions (>60%) in aqueous humor pro-inflammatory cytokine concentrations (IL-6, TNF-α), effectively mitigating cellular infiltration in both ciliary body and iris tissues(Sen H N et al, 2024; Brar, V.S et al, 2024). Secondly, the preoperative long-term use of corticosteroid eye drops in this patient population may mitigate postoperative fibrosis of the filtering bleb (Honjo M et al, 2021; Dave B et al, 2024). Thirdly, the intraoperative use of antimetabolite agents (including mitomycin C) demonstrates threefold therapeutic effects: fibroblast proliferation inhibition, postoperative bleb scarring risk reduction, and establishment of durable intraocular pressure regulation (Weinreb et al.,2016). In the other group, because diagnostic review revealed that 40% of non-uveitis patients had refractory glaucoma including A-R syndrome, neovascular glaucoma, post-traumatic glaucoma, and PACG with previous surgical interventions. According to the previous researches, those refractory cases unexpectedly demonstrated higher scar formation incidence in our cohort analysis, which may be lead to surgery failure (Gedde SJ et al., 2022; Kim M et al., 2015; Mermoud A et al., 1993). These patients necessitate iterative therapeutic interventions for fibrosis control. Overall, our findings indicate that compound trabeculectomy achieves clinically satisfactory outcomes in managing uveitis-associated secondary glaucoma and other glaucoma types, with measurable improvements in key surgical parameters.

For both uveitic and other glaucoma types, compound trabeculectomy achieved similar treatment outcomes with comparable surgery safety. Differences were exclusively observed in complications. Filtering bleb fibrosis and complicated cataract constitute major complications. Success of glaucoma filtering surgery relies on controlling wound healing to prevent sub-conjunctival scarring and maintain bleb functionality. Filtering bleb scarring primarily results from the proliferation of sub-conjunctival fibroblasts and the biosynthesis of collagen and other extracellular material (Skuta et al, 1987; Costa et al, 1993). Bleb fibrosis manifests clinically through reduced height (low to flat), sustained angry vascularization, and corkscrew-shaped or straightened conjunctival vessels over the bleb (Collignon et al, 2005). The IOP may be normal at this stage, therefore recognizing these early signs of failure is critical for timely intervention. Bleb needling, a minimally invasive extraocular intervention for scarred blebs, follows a subconjunctival injection (5–10 mm from the bleb) of 0.5 mL 5 - FU (10 mg/mL) or 0.1 mL MMC (0.2/0.4 mg/mL dilution, total 0.04 mg) by 15 to 20 minutes (Pathak-Ray et al, 2018). Completion requires both lysis of fibrous adhesions and digital palpation confirmation of sufficient ocular softening. Comparative studies on needling with MMC vs 5 - FU in Asian eyes report success rates of 57% vs 48% respectively (Pathak-Ray et al, 2018; Tsai et al, 2015). The application of antibiotic-steroid combination drops is 4 to 6 times daily for one week, and then switch to steroid-only regimen and gradually taper over 2 or 3 weeks. Regular bleb massage should begin from postoperative day 3 to facilitate aqueous drainage. Close monitoring of IOP changes is essential, with pressure below 10mmHg within the first week usually indicating favorable long-term outcomes (Mardelli et al, 1996). Anterior segment OCT can be used to evaluate changes in the number of microcysts in the bleb. Future developments to improve bleb longevity encompass several innovative approaches. Targeted molecular therapies represent a significant direction, such as anti-TGF-β monoclonal antibodies like Lerdelimnumb and YAP/TAZ inhibitors such as Verteporfin (Shao et al, 2023). Novel antifibrotic agents show great promise, exemplified by 0.5% pirfenidone eye drops and LOX antibodies (Shao et al, 2023; Fan Gaskin et al, 2014). Advanced drug delivery systems under development include LDL-chitosan nanoparticles loaded with MMC and biodegradable PLGA implants that enable sustained release of antifibrotic drugs (Fan Gaskin et al, 2014; Kavitha et al, 2024).

Postoperative cataract incidence in the uveitis group demonstrated a twofold increase compared to the non-uveitis cohort. Existing evidence confirms significantly higher rates of complicated cataract formation following trabeculectomy in uveitic secondary glaucoma versus other glaucoma subtypes. Although non-UG eyes also often encounter cataract progression after trabeculectomy, previous clinical studies reported the high frequency of postoperative cataract progression in UG eyes (28.3% to 51.6%) (Lazaro et al, 2002; Husain et al, 2006; Kaburaki et al,2009; Ceballos et al,2002). The observed disparity originates from the unique pathophysiology of uveitis: (a) chronic intraocular inflammation triggers lens epithelial dysfunction through elevated aqueous humor concentrations of IL-6 and TNF-α, which induce oxidative stress damage to lens cells (Pham et al., 2023; Sen et al., 2024; Nema et al., 2018). (b) postoperative inflammatory recurrence establishes cumulative oxidative stress cycles, progressively accelerating lens opacification (Kaburaki et al., 2009). Concurrently, surgery procedure potentiates lens opacification via inflammatory pathways: (i) surgical intervention induces transient inflammatory responses marked by prostaglandin E2 and leukotriene B4 surges, compromising blood-aqueous barrier integrity and lens capsule permeability (Nema et al., 2018; Girkin et al., 2004). (ii) prolonged corticosteroid regimens for inflammation management impair lens epithelial mitochondrial function while depleting antioxidant reserves, thereby potentiating oxidative injury (Lee et al., 2011; Kaburaki et al., 2009).

The pathogenesis of uveitis-associated choroidal detachment demonstrates distinct immunopathological characteristics, despite its low incidence (n=1 in this cohort). Chronic intraocular inflammation mediates Th17/Treg axis imbalance and Th1 hyperactivation, driving IL-6, TNF-α, and VEGF overproduction (Yanoff et al., 2009; Nema et al., 2018; Sen et al., 2024). These cytokines disrupt choroidal vascular integrity through blood-retinal barrier breakdown, manifesting as exudative choroiditis with stromal edema and inflammatory infiltration (Nema et al., 2018; Sen et al., 2024). Mitochondrial impairment exacerbated oxidative stress via reactive oxygen species (ROS) overproduction and NLRP3 inflammasome signaling activation, as demonstrated in recent mechanistic studies (Hered et al., 2024; Belfort et al., 2024). In non-uveitis patients, prolonged elevated intraocular pressure compromises choroidal vascular autoregulation, while abrupt postoperative pressure reduction induces hydrodynamic instability in the suprachoroidal space, resulting in serous effusion (Quintero et al., 2022; Choplin et al., 2014). Clinical management of these pressure-induced detachments generally requires only medical therapy, demonstrating favorable resolution without surgical treatment. Meanwhile, in our non-uveitis cohort, one malignant glaucoma case emerged in a PACG patient with prior filtration surgery, where surgical recurrence-induced fibrotic tissue remodeling increased ciliary block risk, amplifying PACG-specific postoperative vulnerability (Jin et al., 2023). One neovascular glaucoma (NVG) case developed hyphema due to inherent fibrovascular membrane instability, a recognized surgical risk factor. The absence of endophthalmitis and suprachoroidal hemorrhage in our cohort likely reflects stringent surgical protocols and perioperative care optimization (Weinreb et al., 2016).

This study is constrained by its single-center design, limited sample size, and lack of etiological stratification across uveitis subtypes. Future investigations will employ multi-center prospective randomized controlled trials with subgroup analyses stratified by uveitis classification and inflammatory activity, thereby optimizing evidence-based surgical criteria for compound trabeculectomy.

## Conclusion

Compound trabeculectomy manifests suitable for surgically indicated uveitic glaucoma and long-term safety and efficacy. This surgery can effectively lower intraocular pressure, reduce medication dependence and complication rates of uveitic glaucoma patients. Success rates between uveitic and non-uveitic glaucoma cases remain comparable. Vigilance against bleb fibrosis and cataract progression remains essential for both cohorts.

## Data Availability

All data produced in the present study are available upon reasonable request to the authors.

## Reference

[1] Bloch-Michel E, Nussenblatt RB. International Uveitis Study Group recommendations for the evaluation of intraocular inflammatory disease. Am J Ophthalmol. 1987;103(2):234–235. doi:10.1016/s0002-9394(14)74235-7

[2] Massa H, Pipis SY, Adewoyin T, Vergados A, Patra S, Panos GD. Macular edema associated with non-infectious uveitis: pathophysiology, etiology, prevalence, impact and management challenges. Clin Ophthalmol. 2019;13:1761–1777. Published 2019 Sep 10. doi:10.2147/OPTH.S180580

[3] Sudharshan S, Ganesh SK, Biswas J. Current approach in the diagnosis and management of posterior uveitis. Indian J Ophthalmol. 2010;58(1):29–43. doi:10.4103/0301-4738.58470

[4] Yanoff M, Duker J S. Ophthalmology[M]. Elsevier Health Sciences, 2009.

[5] Yang J, Jiang J, Zhang S, et al. Analysis of Posner-Schlossman syndrome in Wenzhou district over the last 10 years [published online March 1, 2017]. Zhongguo Shiyong Yanke Zazhi [Chinese Journal of Practical Ophthalmology]. 2017;35(3):339–342. doi:10.3760/cma.j.issn.1006-4443.2017.03.027

[6] Pan XJ. Who Will Win In The Battle Against The Virus? [Conference presentation]. The Fourth Zhejiang Provincial Forum on Glaucoma Prevention and Control at the Grassroots Level, Hangzhou, China. August 22, 2020case-control.

[7] Škrlová E, Svozílková P, Heissigerová J, Fichtl M. PATHOGENESIS AND CURRENT METHODS OF TREATMENT OF SECONDARY UVEITIC GLAUCOMA. A REVIEW. ETIOPATOGENEZE SOUč ASNÝCH MOŽNOSTí TERAPIE SEKUNDÁRNíHO UVEITICKÉHO GLAUKOMU. PŘEHLED. Cesk Slov Oftalmol. 2023;79(3):111–115. doi:10.31348/2023/7

[8] Pillai MR, Balasubramaniam N, Wala N, et al. Glaucoma in Uveitic Eyes: Long-Term Clinical Course and Management Measures. Ocul Immunol Inflamm. 2024;32(6):1041–1047. doi:10.1080/09273948.2023.2202740

[9] Rojas-Carabali W, Mejía-Salgado G, Cifuentes-González C, Chacón-Zambrano D, Cruz-Reyes DL, Delgado MF, Gómez-Goyeneche HF, Saad-Brahim K, de-la-Torre A. Prevalence and clinical characteristics of uveitic glaucoma: multicentric study in Bogot á, Colombia. Eye (Lond). 2024 Mar;38(4):714–722. doi: 10.1038/s41433-023-02757-9

[10] Nema, H., Nema, N. Gems of Ophthalmology: Glaucoma[M]. Jaypee Brothers Medical Publishers Pvt. Limited, 2018

[11] Koktekir BE, Gedik S, Bakbak B. Bilateral severe anterior uveitis afterunilateral selective laser trabeculoplasty _ J J. Clin Exp Ophthalmol 2013, 41 (3):305–307.

[12] Ma T, Sims JL, Bennett S, et al. High rate of conversion from ocular hypertension to glaucoma in subjects with uveitis. Br J Ophthalmol. 2022 Nov;106(11):1520–1523

[13] Modi N, Vahdani K, Booth AP. Glaucoma surgery. J Perioper Pract. 2011;21(1):33–37. doi:10.1177/175045891102100105

[14] Urbonavičiūtė D, Buteikienė D, Janulevičienė I. A Review of Neovascular Glaucoma: Etiology, Pathogenesis, Diagnosis, and Treatment. Medicina (Kaunas). 2022;58(12):1870.

[15] Chinese Glaucoma Society, Ophthalmology Society of Chinese Medical Association. Expert Consensus on Compound Trabeculectomy in China (2017) [in Chinese]. Zhonghua Yan Ke Za Zhi. 2017;53(4):249–251. doi:10.3760/cma.j.issn.0412-4081.2017.04.004

[16] Gao Y, Zhao Q, Li H, Li J, Li P. Peripheral iridectomy for glaucoma is more effective than compound trabeculectomy and significantly reduces Hcy and hs-CRP levels. Am J Transl Res. 2022;14(10):7451–7458. Published 2022 Oct 15.

[17] Wang Q, Zeng W, Zeng W, Liu Y, Ke M. Clinical Differences between Posner-Schlossman Syndrome Patients with Intermittent Intraocular Pressure Elevation and Glaucomatous Damage. Ophthalmic Res. 2023;66(1):1198–1205. doi:10.1159/000533495

[18] Yuan J, Wang Y, Wang P, Ma R. Safety in Treating Primary Acute Angle-closure Glaucoma under High Intraocular Pressure by Compound Trabeculectomy. Pak J Pharm Sci. 2016;29(6 Spec):2217–2220.

[19] Kanaya R, Kijima R, Shinmei Y, et al. Surgical Outcomes of Trabeculectomy in Uveitic Glaucoma: A Long-Term, Single-Center, Retrospective Case-Control Study. J Ophthalmol. 2021;2021:5550776. Published 2021 May 21. doi:10.1155/2021/5550776

[20] Kaburaki T, Koshino T, Kawashima H, et al. Initial trabeculectomy with mitomycin C in eyes with uveitic glaucoma with inactive uveitis. Eye (Lond). 2009;23(7):1509–1517. doi:10.1038/eye.2009.117-cme

[21] Iwao K, Inatani M, Seto T, et al. Long-term outcomes and prognostic factors for trabeculectomy with mitomycin C in eyes with uveitic glaucoma: a retrospective cohort study. J Glaucoma. 2014;23(2):88–94. doi:10.1097/IJG.0b013e3182685167

[22] Gao X, Lv A, Lin F, et al. Efficacy and safety of trabeculectomy versus peripheral iridectomy plus goniotomy in advanced primary angle-closure glaucoma: study protocol for a multicentre, non-inferiority, randomised controlled trial (the TVG study). BMJ Open. 2022;12(7):e062441. Published 2022 Jul 4. doi:10.1136/bmjopen-2022-062441

[23] Zhang X, Lin F, Li F, Lee JWY, Tham CC. Minimally Invasive Glaucoma Surgery: A New Era in Glaucoma Treatment. Asia Pac J Ophthalmol (Phila). 2023;12(6):509–511. doi:10.1097/APO.0000000000000648

[24] Weinreb, R. N., Grajewski, A. L., Papadopoulos, M., Grigg, J., & Freedman, S. (Eds.). Childhood glaucoma[M]. Kugler Publications, 2013.

[25] Weinreb, R. N.,Crowston J G. Glaucoma Surgery-Open Angle Glaucoma[M]. Kugler Publications, 2016.

[26] Zhou M, Wang W, Huang W, Zhang X. Trabeculectomy with versus without releasable sutures for glaucoma: a meta-analysis of randomized controlled trials. BMC Ophthalmol. 2014;14:41. Published 2014 Mar 31. doi:10.1186/1471-2415-14-41

[27] Kwon HJ, Kong YXG, Tao LW, et al. Surgical outcomes of trabeculectomy and glaucoma drainage implant for uveitic glaucoma and relationship with uveitis activity. Clin Exp Ophthalmol. 2017;45(5):472–480. doi:10.1111/ceo.12916

[28] Feldman RM, Bell NP, eds. Complications of Glaucoma Surgery. New York, NY: Oxford University Press; 2013.

[29] Kojima S, Inoue T, Nakashima K, Fukushima A, Tanihara H. Filtering blebs using 3-dimensional anterior-segment optical coherence tomography: a prospective investigation. JAMA Ophthalmol. 2015;133(2):148–156. doi:10.1001/jamaophthalmol.2014.4489

[30] Babu K, Konana VK, Ganesh SK, et al. Viral anterior uveitis. Indian J Ophthalmol. 2020;68(9):1764–1773. doi:10.4103/ijo.IJO_928_20

[31] Sen H N. 2024-2025 Basic and Clinical Science Course, Section 9: Uveitis and Ocular Inflammation[M]. American Academy of Ophthalmology, 2024.

[32] Brar, V.S. 2024-2025 Basic and clinical science course, section 2: Fundamentals and principles of ophthalmology[M]. American Academy of Ophthalmology, 2024.

[33] Honjo M, Yamagishi R, Igarashi N, et al. Effect of postoperative corticosteroids on surgical outcome and aqueous autotaxin following combined cataract and microhook ab interno trabeculotomy. Sci Rep. 2021;11(1):747. Published 2021 Jan 12. doi:10.1038/s41598-020-80736-w

[34] Dave B, Patel M, Suresh S, et al. Wound Modulations in Glaucoma Surgery: A Systematic Review. Bioengineering (Basel). 2024;11(5):446. Published 2024 Apr 30. doi:10.3390/bioengineering11050446

[35] Gedde SJ, Feuer WJ, Lim KS, et al. Treatment Outcomes in the Primary Tube Versus Trabeculectomy Study after 5 Years of Follow-up. Ophthalmology. 2022;129(12):1344–1356. doi:10.1016/j.ophtha.2022.07.003

[36] Kim M, Lee C, Payne R, Yue BY, Chang JH, Ying H. Angiogenesis in glaucoma filtration surgery and neovascular glaucoma: A review. Surv Ophthalmol. 2015;60(6):524–535. doi:10.1016/j.survophthal.2015.04.003

[37] Mermoud A, Salmon JF, Straker C, Murray AD. Post-traumatic angle recession glaucoma: a risk factor for bleb failure after trabeculectomy. Br J Ophthalmol. 1993;77(10):631–634. doi:10.1136/bjo.77.10.631

[38] Pham JH, Stankowska DL. Mitochondria-associated endoplasmic reticulum membranes (MAMs) and their role in glaucomatous retinal ganglion cell degeneration-a mini review. Front Neurosci. 2023;17:1198343. Published 2023 May 12. doi:10.3389/fnins.2023.1198343

[39] Skuta GL, Parrish RK 2nd. Wound healing in glaucoma filtering surgery. Surv Ophthalmol. 1987;32(3):149–170. doi:10.1016/0039-6257(87)90091-9

[40] Costa VP, Spaeth GL, Eiferman RA, Orengo-Nania S. Wound healing modulation in glaucoma filtration surgery. Ophthalmic Surg. 1993;24(3):152–170.

[41] Collignon NJ. Wound healing after glaucoma surgery: how to manage it?. Bull Soc Belge Ophtalmol. 2005;(295):55–59.

[42] Pathak-Ray V, Choudhari N. Rescue of failing or failed trabeculectomy blebs with slit-lamp needling and adjunctive mitomycin C in Indian eyes. Indian J Ophthalmol. 2018;66(1):71–76. doi:10.4103/ijo.IJO_523_17

[43] Tsai AS, Boey PY, Htoon HM, Wong TT. Bleb needling outcomes for failed trabeculectomy blebs in Asian eyes: a 2-year follow up. Int J Ophthalmol. 2015;8(4):748–753. Published 2015 Aug 18. doi:10.3980/j.issn.2222-3959.2015.04.19

[44] Mardelli PG, Lederer CM Jr, Murray PL, Pastor SA, Hassanein KM. Slit-lamp needle revision of failed filtering blebs using mitomycin C. Ophthalmology. 1996;103(11):1946–1955. doi:10.1016/s0161-6420(96)30403-x

[45] Shao CG, Sinha NR, Mohan RR, Webel AD. Novel Therapies for the Prevention of Fibrosis in Glaucoma Filtration Surgery. Biomedicines. 2023;11(3):657. Published 2023 Feb 21. doi:10.3390/biomedicines11030657

[46] Fan Gaskin JC, Nguyen DQ, Soon Ang G, O’Connor J, Crowston JG. Wound Healing Modulation in Glaucoma Filtration Surgery-Conventional Practices and New Perspectives: The Role of Antifibrotic Agents (Part I). J Curr Glaucoma Pract. 2014;8(2):37–45. doi:10.5005/jp-journals-10008-1159

[47] Kavitha S, Tejaswini SU, Venkatesh R, Zebardast N. Wound modulation in glaucoma surgery: The role of anti-scarring agents. Indian J Ophthalmol. 2024;72(3):320–327. doi:10.4103/IJO.IJO_2013_23

[48] Lazaro C, Benitez-del-Castillo JM, Castillo A, et al. Lens fluorophotometry after trabeculectomy in primary open-angle glaucoma. Ophthalmology. 2002;109:76–79.

[49] Husain R, Aung T, Gazzard G, et al. Effect of trabeculectomy on lens opacities in an east Asian population. Arch Ophthalmol. 2006;124:787–792.

[50] Ceballos EM, Beck AD, Lynn MJ. Trabeculectomy with antiproliferative agents in uveitic glaucoma. J Glaucoma. 2002;11(3):189–196. doi:10.1097/00061198-200206000-00005

[51] Girkin C. Glaucoma: Science and practice[J]. 2004.

[52] Lee S, Van Bergen NJ, Kong GY, et al. Mitochondrial dysfunction in glaucoma and emerging bioenergetic therapies. Exp Eye Res. 2011;93(2):204–212. doi:10.1016/j.exer.2010.07.015

[53] Hered R W. 2024-2025 Basic and Clinical Science Course, Section 6: Pediatric Ophthalmologie and Strabismus[M]. American Academy of Ophthalmology, 2024.

[54] Belfort R N, Singh A D. 2024-2025 Basic and Clinical Science Course, section 4: ophthalmic pathology and intraocular tumors[M]. American Academy of Ophthalmology, 2024.

[55] Quintero H, Shiga Y, Belforte N, et al. Restoration of mitochondria axonal transport by adaptor Disc1 supplementation prevents neurodegeneration and rescues visual function. Cell Rep. 2022;40(11):111324. doi:10.1016/j.celrep.2022.111324

[56] Choplin, N. T., & Traverso, C. E. (Eds.). Atlas of glaucoma[M]. CRC Press, 2014.

[57] Jin SW, Caprioli J. Long-term Treatment Outcomes for Malignant Glaucoma. Ophthalmol Glaucoma. 2024;7(3):282–289. doi:10.1016/j.ogla.2023.12.005

